# Projected long-term impacts of US funding cuts on TB and HIV in South Africa and the TB Programme response

**DOI:** 10.64898/2026.03.25.26349165

**Authors:** Mmamapudi Kubjane, Lise Jamieson, Leigh F. Johnson, Jody Boffa, Priashni Subrayen, Fareed Abdullah, Norbert Ndjeka, Limakatso Lebina, Pren Naidoo, Erika Mohr-Holland, Gesine Meyer-Rath

## Abstract

**Background:** Reductions in United States (US) funding for tuberculosis (TB) and HIV programmes have raised concerns and challenges for TB care and management in South Africa and globally. We used mathematical modelling to illustrate the potential impact of unmitigated funding disruptions, estimating long-term effects on TB incidence and mortality over 2025–2035.

**Methods:** Stakeholder-informed scenarios were modelled, assuming both minimal and maximal disruptions to key TB and HIV services, including preventive therapy for people living with HIV (PLHIV), TB testing, TB treatment initiation, and antiretroviral therapy (ART) coverage.

**Results and discussion:** Between 2025 and 2035, reduced ART coverage was projected to result in 235,000–1,000,000 additional HIV infections; and a 12–41% increase in TB episodes, and 21–72% rise in TB deaths among people living with HIV. Overall, 220,000–730,000 additional TB episodes and 67,000–225,000 TB deaths are anticipated, potentially reversing years of progress. Although mitigation efforts were not included in the model, early responses in South Africa have involved increased diagnostic testing and additional domestic funding. To maintain progress in TB and HIV control, rapid and sustained programmatic responses and funding are essential to prevent substantial setbacks and avert avoidable illness and death.

## Introduction

South Africa has made great strides in reducing TB, with TB incidence and mortality dropping by 30.4% and 47.7% respectively between 2009 and 2019, primarily driven by expanded testing and antiretroviral therapy (ART) scale-up [1]. Despite these gains, the country carries one of the highest burdens of TB and TB-HIV co-infection worldwide. In 2024, we estimated approximately 320,000 new TB episodes and 62,000 deaths, half occurring among people living with HIV (PLHIV) [2].

The momentum of these gains were threatened by the US government’s withdrawal of funding in early 2025. In 2024/25, South Africa’s National TB Programme (NTP) was largely domestically funded (67%), with external funding from the Global Fund (19%), and the USAID and PEPFAR (14%); while PEPFAR funded 14% of the HIV programme [5].

Following our report on the immediate impacts of US funding cuts on TB services and research in South Africa [6], the NTP tasked us with estimating the potential long-term consequences for TB incidence and mortality under scenarios in which these funding reductions are unmitigated. Additionally, we provide an update on the NTP’s response to the funding cuts. This report aims to inform urgent policy and financing decisions by quantifying the potential long-term consequences of service disruptions and highlighting the scale of the remaining recovery efforts required to prevent the reversal of hard-won gains in TB prevention and care.

## Methods

We used the Thembisa TB/HIV mathematical model, which dynamically combines TB programme interventions (preventive therapy, testing, and treatment) with HIV epidemic dynamics and interventions (HIV prevention, testing, and ART) [1,2]. We modelled intervention reduction scenarios informed by NTP stakeholder consultations [7], providing plausible intervention reduction ranges as minimum and maximum scenarios (Table 1), which were triangulated with expenditure data [4]. Scenarios assumed varying reductions in TB preventive therapy, testing, and treatment linkage. HIV service scenarios varied ART coverage only, given its strong effect in reducing progression to active TB disease and TB-related deaths among PLHIV [8].

**Table 1:** Stakeholder-informed assumptions regarding reductions in TB and HIV programme intervention coverages.

|  | Minimum | Maximum |
| --- | --- | --- |
| **Baseline** | - | - |
| **TB programme interventions** |  |  |
| TB preventive therapy for people living with HIV | 20% | 25% |
| TB testing and screening (outreach, finding vulnerable clients, collecting sputum samples) | 20% | 25% |
| Linkage to TB treatment | 15% | 25% |
| **HIV programme interventions** |  |  |
| Antiretroviral treatment coverage | 14% | 28% |

We estimated additional TB cases and deaths under four scenarios without additional resources to close the US funding gap: minimum–maximum reductions without recovery (2025–2035); and minimum–maximum reductions with gradual recovery to baseline levels starting in 2029, corresponding to the end of the current US administration’s term.

## Results

ART coverage reductions of 14–28% alone increased new HIV infections (2025–2035) by 235,000 (16%) with recovery, and up to 1,000,000 (67%) without recovery (Figure 1A). The resulting higher HIV incidence and lower ART coverage drove a 12–41% rise in HIV-associated TB episodes and increase in 21–72% deaths among PLHIV.

**Figure 1:**
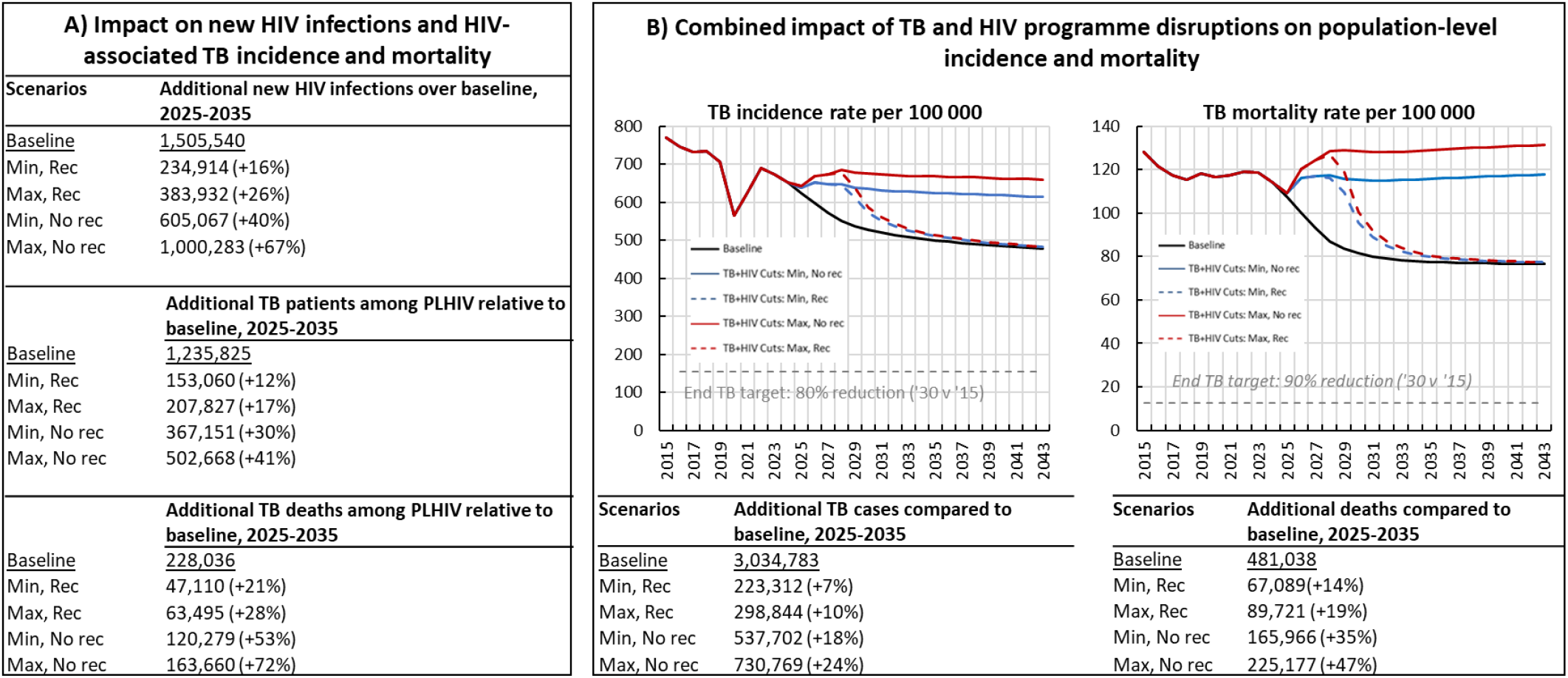
Projected increases in HIV incidence, HIV-associated TB, and overall TB incidence and mortality, 2025-2035.

We projected additional TB cases ranging from 223,000 (7%) in the ‘minimal impact with recovery’ scenario to 731,000 (24%) in the ‘maximal impact with no recovery’ scenario (Figure 1B). The mortality impact would be even more severe: relative to the baseline estimate of 481,000 TB deaths between 2025 and 2035, additional deaths could range from 67,000 (14%) in the ‘minimal impact with-recovery’ scenario to 225,000 (47%) in the ‘maximal impact without recovery’ scenario (Figure 1B). Recovery scenarios would require an additional 8–12 years to return TB incidence and mortality to baseline trajectories, highlighting the long-lasting consequences of even temporary service disruptions.

### National TB Programme Response

The South African NTP has responded to mitigate the impact of these funding cessations through annual TB Recovery Plans that prioritised TB diagnosis, linkage and retention in care [9,10]. One of the key priorities of the 2025/26 TB Recovery Plan, linked to the End TB Campaign, was to test 5 million people for TB. During the 2025 calendar year, approximately

3.56 million tests were conducted [10]. Although testing reached only 71% of the target, the campaign likely offset the projected 20–25% (600,000–800,000) reduction in testing capacity under the modelled funding disruption scenarios.

Furthermore, in 2025, the South African government announced an allocation of an additional US$ 41.7 million in emergency funds to mitigate the impact of the withdrawal of PEPFAR funding [11,12]. Of this amount, US$ 32 million has been allocated to provinces through the District Health Program grant for comprehensive HIV services, and US$ 7.1 million to the South African Medical Research Council for HIV research. However, the specific allocation to TB programmes is unclear.

## Discussion

We updated our initial report [6], modelling the long-term impacts of unmitigated funding disruptions, which could have catastrophic consequences on TB incidence and mortality. In the worst-case scenario, without service recovery, there could be an additional 731,000 TB cases and 225,000 deaths, erasing more than a decade of progress. Even with complete service restoration in 2029, it could take 8–12 years to return to baseline trends. Highlighting that delayed responses to these funding disruptions could have long-lasting consequences, amplifying TB illness and lives lost. This is because when highly prevalent TB is undetected and untreated TB spreads rapidly, requiring years of sustained testing, treatment, and financial investment to reverse.

Projections include a 12–41% increase in additional cases and 21%–72% in TB deaths among PLHIV, highlighting the integrated nature of the TB and HIV epidemics in this high-burden setting. It also demonstrates how disruptions to HIV services have profound consequences for TB. ART scale-up remains one of the most effective interventions for preventing TB in South Africa [8], and any reversal of ART coverage will unsurprisingly drive increases in HIV-associated TB and the overall burden of disease.

The South African government’s allocation of US$ 41.7 million [11,12] to mitigate the funding gaps represents an important step towards domestic resource mobilisation. However, transparency on how much of these funds are ring-fenced for TB services is required to avoid neglecting the TB programme and to strengthen accountability. Additionally, the current funding crisis presents an opportunity for the TB community to explore alternative financing mechanisms, such as social impact bonds [13].

The End TB Campaign [10] demonstrates what can be possibly achieved with coordinated collective action and political will. However, for sustained long term impact, efficient and strategic implementation is required. Innovations such as the near-point-of-care PlusLife MiniDock MTB test, can reduce diagnostic costs. In the longer term, effective adult vaccines may provide reduce the TB burden further, though will come at an additional cost. Their impact will depend on coordinated planning, financing, and integration into adequately functioning TB and HIV programmes.

Given the NTP’s additional funding and increased testing, our projections of the impact of US funding cuts are pessimistic. Nonetheless, these results are important in illustrating potential impact without of mitigation efforts. Sustained political commitment, transparent resource allocation, and protection of core TB and HIV services remain essential to prevent reversals in progress toward TB elimination.

## Data Availability

All data produced in the present study are available upon reasonable request to the authors

